# Endothelial Activation and Stress Index Predicts Mortality in Critically Ill Atrial Fibrillation Patients: A Retrospective Cohort Analysis Using the MIMIC-IV Database

**DOI:** 10.1101/2025.05.29.25328606

**Authors:** Mei-mei Li, Ping-yu Cai, Nian Li, Jinxia Liu, Yaoxin Ruan, Jiaxiang Pan, Fengjing Cai, Chaoyang Xu, Hui-li Lin

## Abstract

**Background:** The Endothelial Activation and Stress Index (EASIX) reflects endothelial dysfunction and has prognostic value in cardiovascular diseases. This study investigates the association between EASIX and all-cause mortality in critically ill atrial fibrillation (AF) patients.

**Methods:** Using MIMIC-IV (v3.1) data, 4722 AF patients were stratified by EASIX quartiles. Cox regression, Kaplan-Meier curves, and restricted cubic spline (RCS) models assessed mortality risk at 7, 30, 180, and 365 days. Subgroup analyses evaluated consistency.

**Results:** EASIX was significantly associated with increased mortality at all time points. The Kaplan-Meier survival curve shows that as the EASIX quartile increases, the 7-day, 30-day, 180-day, and 365-day mortality rates of AF patients significantly increase. RCS analyses showed that there was a significant non-linear relationship between EASIX and the 7-day, 30-day, 180-day, and 365-day mortality rates of patients with AF. The Cox analysis before and after adjusting for confounding factors showed that EASIX was an independent risk factor for 7-day, 30-day, 180-day, and 365-day all-cause mortality in AF patients. Subgroup analyses further indicated the robustness of these results.

**Conclusions:** EASIX levels are significantly correlated with all-cause mortality at 7, 30, 180, and 365 days in patients with AF, and endothelial dysfunction plays an important role in poor prognosis in AF patients.

## Introduction

Cardiovascular disease remains one of the leading causes of death worldwide, with AF being one of the most common arrhythmias ^[1]^. Approximately 330 million people worldwide suffer from AF, and its prevalence significantly increases with age ^[2]^. AF is often associated with higher mortality rates, especially in critically ill patients in the intensive care unit (ICU), where the incidence of AF can reach up to 14% ^[3]^.

Endothelial dysfunction is an important factor in the pathogenesis of AF, and its role in AF patients has been confirmed in multiple studies ^[4,5]^. Damage and functional abnormalities of endothelial cells can trigger oxidative stress, release of pro-inflammatory cytokines, and increased vascular permeability, which collectively promote atrial fibrosis and electrical remodeling, further exacerbating the occurrence and maintenance of AF ^[4,6]^. Previous studies have shown that the levels of common serum endothelial markers such as vWF and ICAM-1 are significantly elevated in patients with AF, suggesting that endothelial dysfunction is an important driving force for the development of AF ^[7]^. In addition, the increase in endothelial injury and thrombosis significantly increases the risk of stroke and other cardiovascular adverse events in patients with AF ^[5]^. Therefore, evaluating endothelial function is crucial for predicting and managing clinical outcomes of AF.

EASIX, as a comprehensive indicator reflecting endothelial activation and damage, has unique predictive potential. Compared to other single biomarkers, EASIX combines lactate dehydrogenase (LDH), creatinine, and platelet count, which can more comprehensively reflect the endothelial function status and overall health status of patients ^[8,9]^. Therefore, this study chose EASIX as a predictive indicator for mortality in critically ill AF patients to explore its effectiveness in risk assessment.

## Methods

### Study population

This study utilized data from the Medical Information Mart for Intensive Care, version IV (MIMIC-IV) database, which contains de-identified health-related data from over 40,000 adult patients admitted to the Beth Israel Deaconess Medical Center in Boston, Massachusetts, from 2001 to 2012. This study focused on a study population in MIMIC-IV consisting of 15878 individuals who were diagnosed with AF. Patients aged <18, or with missing data for any of the variables used to calculate the EASIX index, were excluded. Finally, a total of 4,722 participants were included in the final analysis (Figure 1).

**Figure 1.**
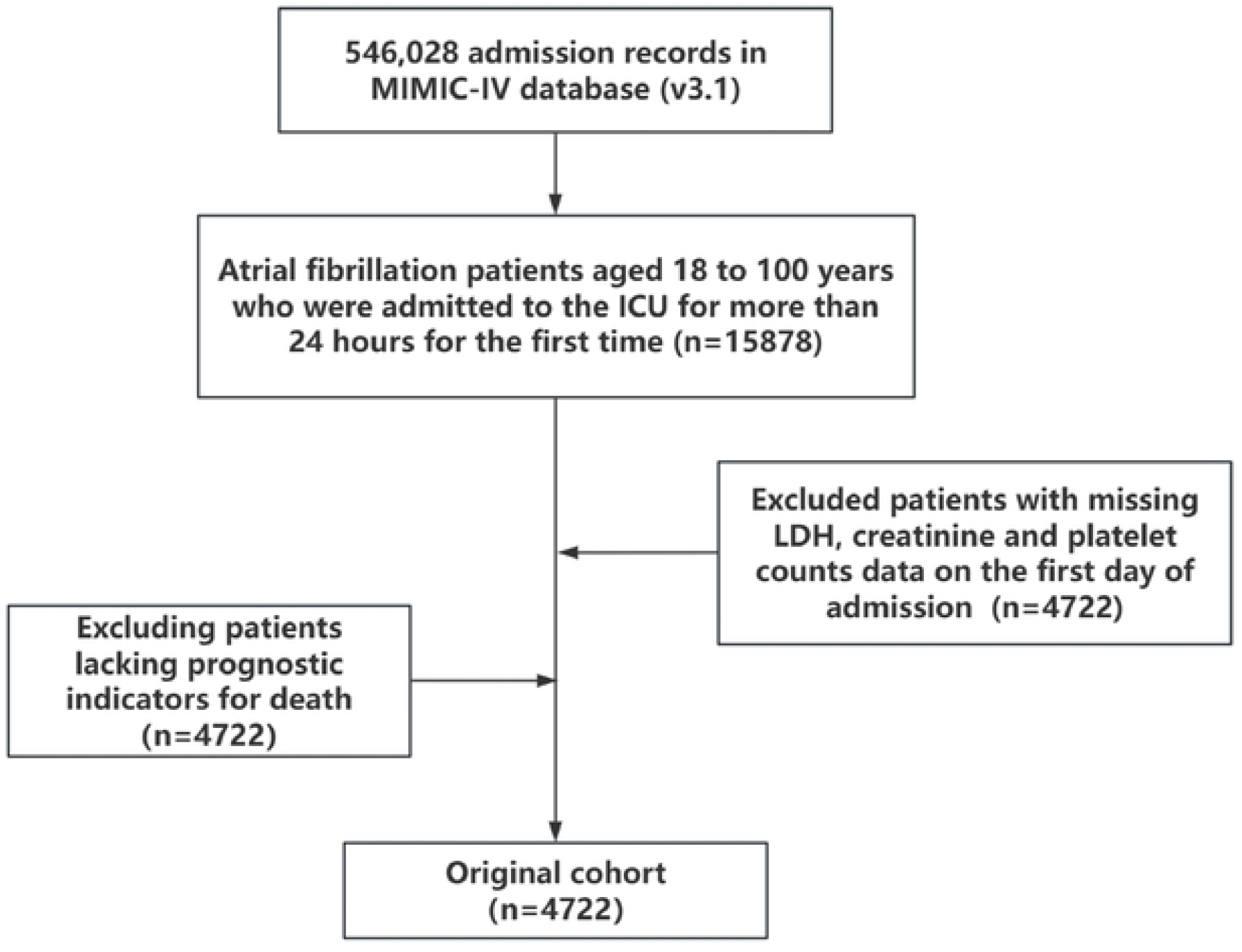
Flow of included patients through the trial.

### Data collection

The pgAdmin PostgreSQL tool (version 6.1) was used to extract information from the MIMIC-IV database, including the primary variable of interest was the EASIX index, calculated as the product of creatinine (mg/dL), platelet count (×10^3^/μL), and lactate dehydrogenase (LDH, U/L). Other characteristics encompassed patient demographics (age, gender and race), vital signs (heart rate (HR), temperature, respiratory rate (RR), peripheral oxygen saturation (Spo2), mean blood pressure (MBP), systolic blood pressure (SBP), and diastolic blood pressure (DBP)), laboratory test results (red blood cell (RBC), white blood cells (WBC), monocytes, neutrophils, hemoglobin, fibrinogen, bilirubin, lymphocytes, platelets, albumin, creatinine, blood urea nitrogen (BUN), apolipoprotein(ALP), alanine aminotransferase(ALT), aspartate transaminase(AST), international normalized ratio(INR), partial thromboplastin time(PTT), prothrombin time(PT), potassium, sodium, calcium, glucose, and LDH) within the initial 24 h of ICU admission, as well as clinical outcomes (in-hospital mortality and 7-、30-、180-、365-day mortality). Additionally, various complications (including hypertension, diabetes, myocardial infarct, congestive heart failure, cerebrovascular disease, peripheral vascular disease, dementia, chronic pulmonary disease, rheumatic disease, paraplegia, malignant cancer, severe liver disease and peptic ulcer disease), and the charlson comorbidity index were included as indicators to comprehensively assess the patients’ overall clinical status.

### Statistical Analysis

Patients were divided into 4 groups according to the optimal cut-off value of EASIX. All data were presented as mean ± standard deviation or median (interquartile range) for continuous variables and counts (percentage) for categorical variables, respectively. Continuous variables were compared by students ‘t-test (normally distributed) or Mann-Whitney U-test (non-normally distributed). Categorical variables were analyzed using chi-square test or Fisher’s exact test. Cox proportional hazards model was used to evaluate the association between the EASIX index and mortality. Kaplan-Meier was performed survival analysis to assess the relationship between different quartiles of the EASIX index and survival outcomes. To assess the discriminative ability of the EASIX index, we plotted the receiver operating characteristic (ROC) curve for 7-、30-、180- and 365-day mortality. Subgroup analyses were performed to evaluate whether the predictive power of the EASIX index differed across various subgroups defined by age, sex, and comorbidities. Restricted cubic spline curve was used to evaluate the correlation between continuous EASIX and mortality. Statistical analyses were conducted using the R statistical language (version 4.2.1; R Foundation, Vienna, Austria) and SPSS (version 26.0; IBM, Armonk, New York, United States). P<0.05 (two-tailed) was considered statistically significant.

## Results

### Baseline characteristics

The study enrolled a total of 4722 ICU patients with AF, categorized based on EASIX quartiles. Baseline characteristics of participants are detailed in Table 1.

Significant differences were observed across several variables. The mean age showed slight variation, with the youngest group in Q4 (72.81 years, p < 0.001), and females were more prevalent in Q1 compared to Q4 (p < 0.001). Significant race differences were observed (p < 0.001), and clinical parameters such as ALT, AST, creatinine, LDH and BUN were notably higher in Q4 (all p < 0.001), suggesting more severe metabolic and organ dysfunction in this group. Complications, including diabetes (39.88% in Q4, p < 0.001), myocardial infarct (33.02% in Q4, p < 0.001), peripheral vascular disease (16.68% in Q4, p < 0.001), peptic ulcer disease (4.66% in Q4, p =0.027), malignant cancer (20.58% in Q4, p < 0.001) and severe liver disease (10.75% in Q4, p < 0.001) were more prevalent in the higher EASIX groups. Mortality rates significantly increased with higher EASIX quartiles across all time points, with 30-day mortality ranging from 18.04% in Q1 to 41.57% in Q4 (p < 0.001), and 365-day mortality ranging from 35.22% in Q1 to 60.97% in Q4 (p < 0.001). These findings highlight the strong association between higher EASIX scores and worse clinical outcomes, with higher scores reflecting more severe physiological disturbances.

### EASIX and all-cause mortality

Cox regression analysis of 7-day, 30-day, 180-day and 365-day mortality rates, stratified by EASIX quartiles, is shown in Table 2. In Model I, without adjusting for any confounding factors, there was a significant correlation between EASIX score and mortality at all time points for ICU patients with AF. The higher the EASIX value, the higher the risk of mortality at all time points, especially in the Q4 group, where the hazard ratio (HR) was significantly higher than other groups (all groups <0.001). After adjusting for confounding factors, the association between EASIX and mortality remained statistically significant. In an additional analysis, we treated EASIX as a continuous variable in the Cox regression model. The observed trends remained consistent with those obtained using the EASIX quartile classification system, further validating the reliability of EASIX scores in predicting mortality among atrial fibrillation (AF) patients (sTable 1). The receiver operating characteristic curves (sFigure 1) showed that EASIX exhibited moderate predictive ability in predicting 7-day, 30-day, 180-day, and 365-day mortality rates in ICU patients with AF. The AUC values were 0.66, 0.63, 0.61, and 0.61, respectively, indicating stable predictive performance at different time points.

### RCS analyses

RCS analyses in Figure 2 showed that there was an overall non-linear trend relationship between EASIX and the risk of the 7-day, 30-day, 180-day, and 365-day all-cause mortality. Although the overall P-values at all time points were less than 0.001, the non-linear effect P-values for 7-day, 30-day, 180-day and 365-day mortality were 0.274, 0.415, 0.974 and 0.774, respectively, indicating a relatively stable relationship between LnEASIX and mortality at these time points. Moreover, we observed that as LnEASIX increased, the risk of all-cause mortality in patients showed an upward trend.

**Figure 2.**
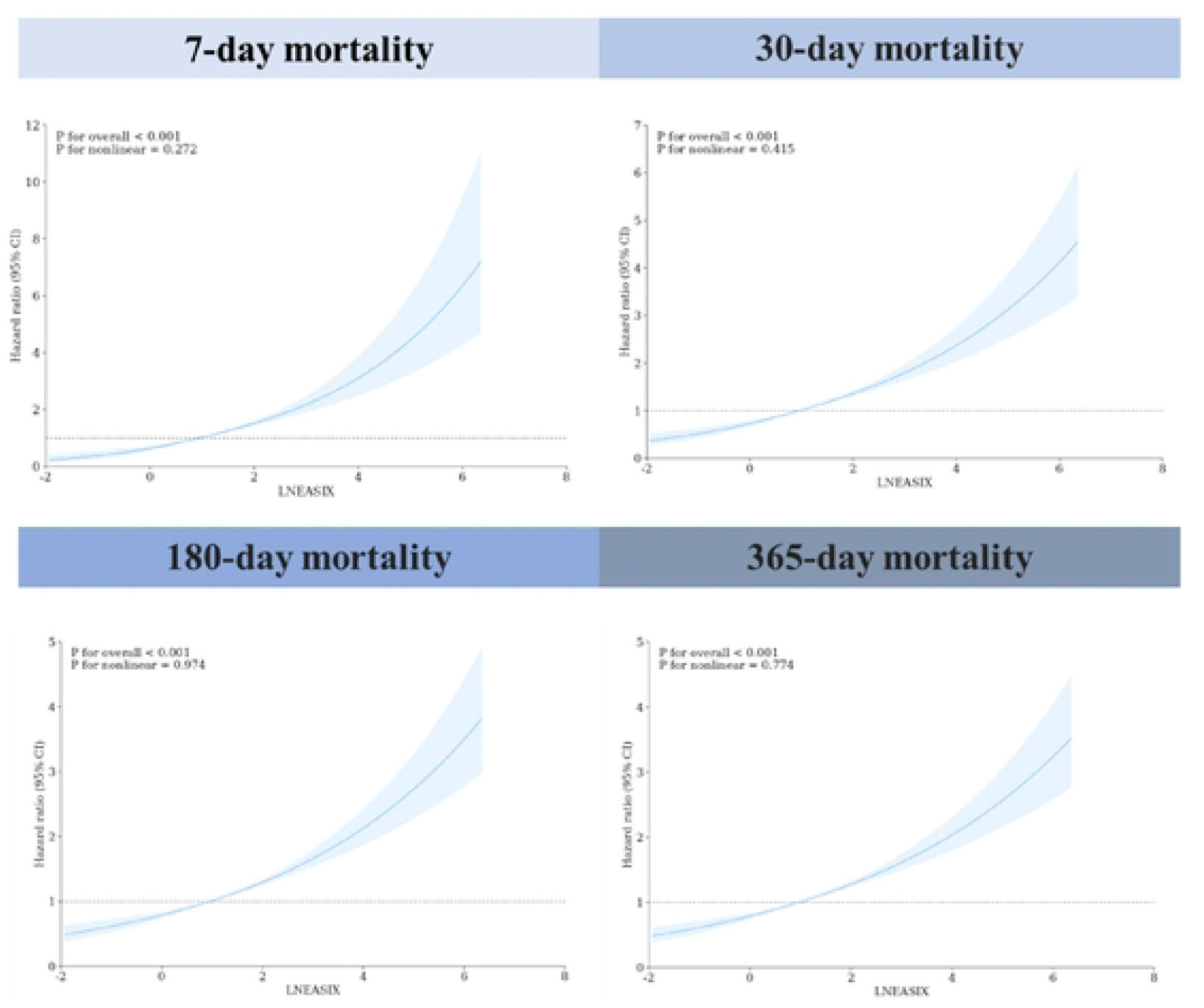
Restricted Cubic Spline Curves for 7-Day, 30-Day, 180-Day and 365-Day mortality in patients with atrial fibrillation

### Kaplan–Meier survival curve analysis

The Kaplan–Meier survival curves, as indicated in Figure 3, showed that as the EASIX quartile increases, the 7-day, 30-day, 180-day, and 365-day mortality rates of AF patients significantly increase. The Log-rank *P*-values at all time points are less than 0.0001, indicating significant survival differences between different EASIX quantile arrays. The high EASIX quantile array (Q4) showed the lowest survival rate, indicating that EASIX can be an effective predictor of mortality risk in patients with AF (Figure 3).

**Figure 3.**
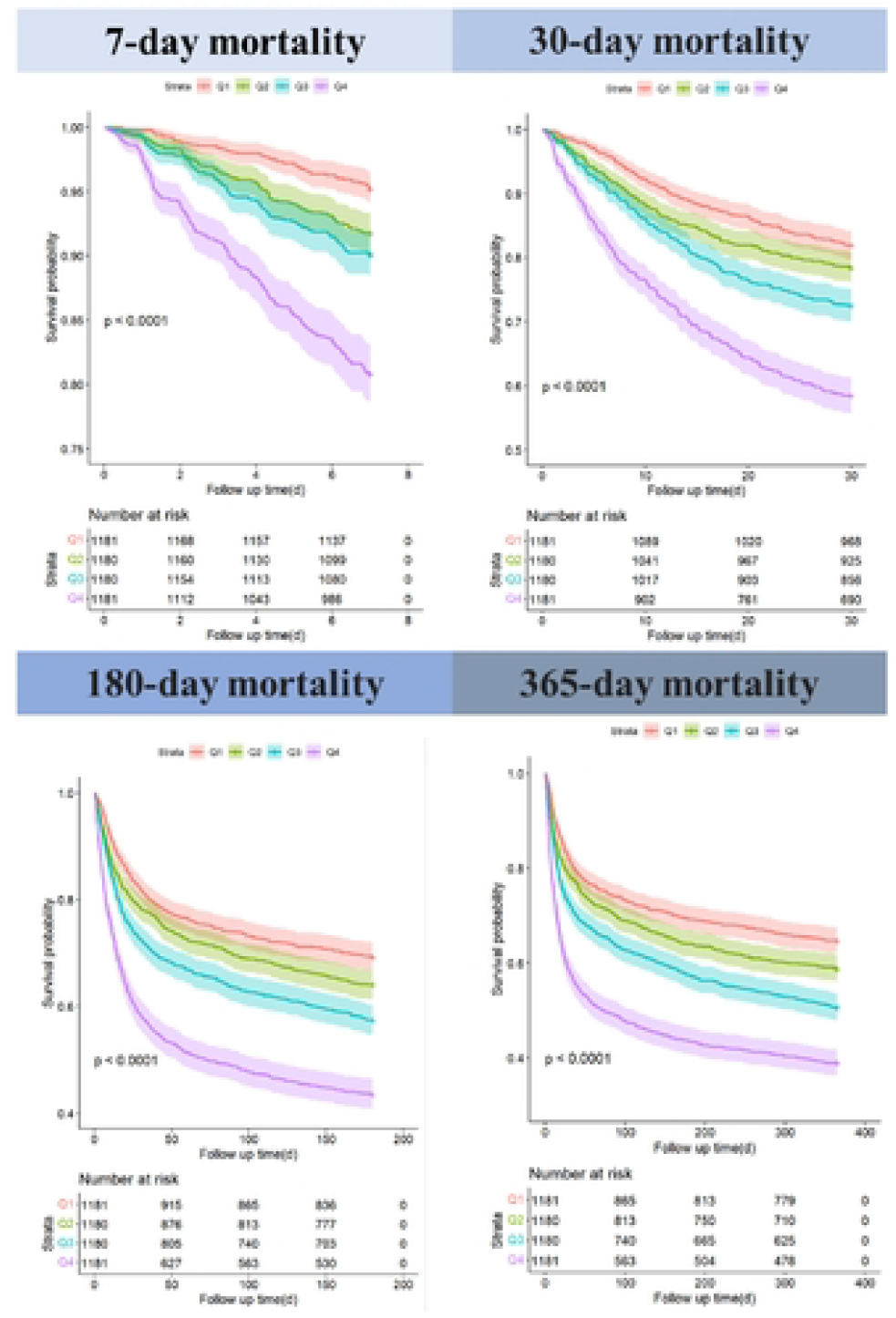
Kaplan-Meier Survival Curves for 7-Day, 30-Day, 180-Day, and 365-Day mortality stratified by EASIX quartiles in patients with atrial fibrillation

### Subgroup analysis

The stability of the association between EASIX and both early and late mortality was evaluated through subgroup analysis (Figure 4). EASIX is significantly correlated with mortality at different age scores, regardless of the time point (7 days, 30 days, 180 days, or 365 days). Similarly, EASIX was significantly positively correlated with 7-day, 30-day, 180-day, and 365-day mortality in both men and women. Whether or not patients had hypertension, diabetes, myocardial infarction, congestive heart failure and cerebrovascular disease, EASIX remained highly correlated with mortality risk, with a significant increase in 7-day, 30-day, 180-days, and 365-day mortality as EASIX levels increased. These findings further reinforce the robustness of the conclusions.

**Figure 4.**
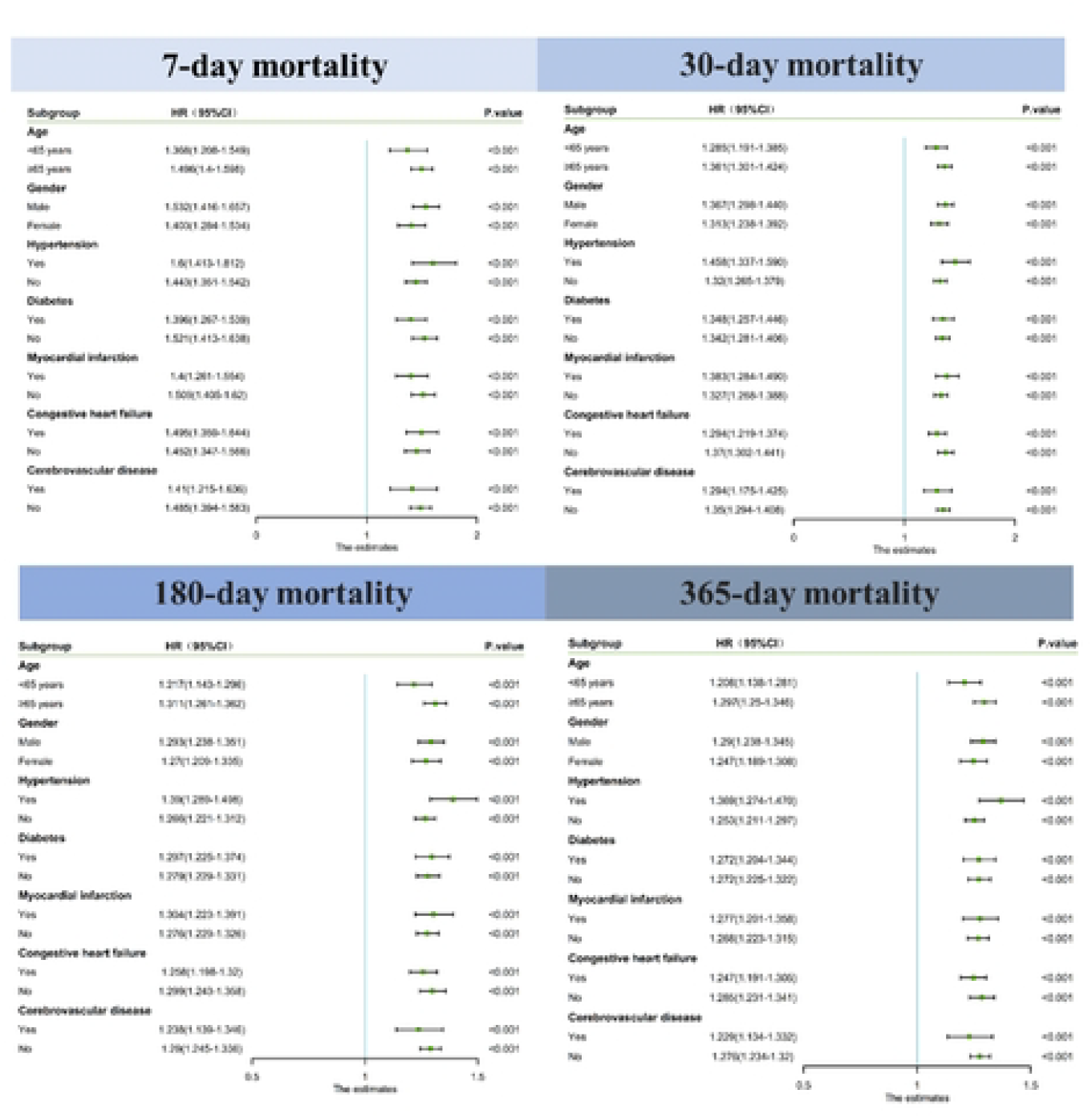
Forest plot for subgroup analysis of the association between EASIX and 7-Day. 30-Day. 180-Day.and 365-Day mortality in patients with atrial fibrillation

## Discussion

This study found that higher levels of EASIX were significantly associated with all-cause mortality in critically ill AF patients, particularly during the 7-day, 30-day, 180-day, and 365-day follow-up periods. The Kaplan-Meier survival curves shows that patients in the highest quartile of EASIX have significantly reduced survival rates at all time points. Multivariate Cox regression analysis also confirmed that patients with higher levels of EASIX have a significantly increased risk of mortality.

Endothelial dysfunction plays an important role in the occurrence and development of AF, which has been increasingly confirmed in clinical and basic research. Dysfunction of endothelial cells can promote the occurrence of AF through various pathways, including oxidative stress, release of pro-inflammatory cytokines, and increased vascular permeability ^[10,11]^. Research has shown that elevated serum endothelial markers such as vWF and ICAM-1 in patients with AF reflect the presence of endothelial dysfunction, which not only exacerbates atrial structure and electrical remodeling, but may also promote thrombosis, significantly increasing the risk of stroke and other adverse events ^[7,12]^. Therefore, assessment and management of endothelial dysfunction are crucial for improving clinical outcomes in patients with AF.

EASIX was initially introduced in allogeneic hematopoietic stem cell transplantation to predict the risk of graft-versus-host disease (GVHD) and non recurrent death. With the deepening of research, EASIX has been applied to the study of other acute and chronic diseases, including infectious diseases and evaluation of multiple organ dysfunction ^[13,14]^. In the field of cardiovascular disease, EASIX has also shown its potential in evaluating endothelial injury and its impact on prognosis, especially in critically ill patients ^[15,16]^. EASIX can comprehensively reflect the status of endothelial activation, oxidative stress, and organ damage, which makes it uniquely advantageous in evaluating the outcomes of cardiovascular disease patients.

This study supplements the value of EASIX as a mortality prediction tool for critically ill AF patients, indicating a significant correlation between higher levels of EASIX and poorer prognosis in patients. Compared with other endothelial function indicators such as vWF, ICAM-1, etc., EASIX combines lactate dehydrogenase, creatinine, and platelet count, which can more comprehensively reflect the endothelial function status and overall health status of patients. This comprehensiveness gives EASIX a significant advantage in risk assessment for patients with fibrillation in critical care units, facilitating earlier risk stratification and more targeted intervention measures.

Although this study demonstrates the potential application value of EASIX in predicting mortality rates in critically ill AF patients, there are still some shortcomings. Firstly, this study is retrospective and there is a possibility of selection bias and confounding factors. Secondly, the research data comes from a single database (MIMIC-IV), and its results may not be applicable to other populations or medical environments. In addition, the relationship between the dynamic changes of EASIX and specific intervention measures is not yet clear, and further prospective studies are needed to verify the relationship between changes in EASIX at different time points and patient prognosis. More multicenter studies are needed in the future to evaluate the generalizability and predictive efficacy of EASIX in different subgroups of AF patients.

## Conclusion

This study emphasizes the potential role of EASIX as a predictor of mortality in critically ill AF patients. The association between elevated EASIX levels and increased all-cause mortality suggests that endothelial dysfunction is a key factor affecting the prognosis of patients with AF. Therefore, EASIX can serve as a valuable tool for early risk stratification and guide treatment strategies to improve prognosis. The potential significance of treating endothelial dysfunction in AF management needs further research and verification.

## Abbreviations

AF: Atrial fibrillation
ALT: Alanine aminotransferase
ALP: Apolipoprotein
AST: Aspartate aminotransferase
AUC: Area under the curve
BUN: Blood urea nitrogen
CA: Cardiac arrest
CI: Confidence interval
DBP: diastolic blood pressure
EASIX: Endothelial Activation and Stress Index
F: Female
GVHD: graft-versus-host disease
HR: Hazard ratios
ICU: Intensive care unit
INR: International normalized ratio
LDH: Lactate dehydrogenase
M: Male
MBP: Mean blood pressure
MIMIC-IV: Medical Information Mart for Intensive Care IV
PT: Prothrombin time
PTT: Partial thromboplastin time
RBC: Red blood cell
ROC: Receiver operating characteristic
RCS: Restricted cubic spline
RR: Respiratory rate
SBP: systolic blood pressure
Spo2: Peripheral oxygen saturation
WBC: White blood cell.

## Acknowledgements

None.

## Authors’ contributions

Meimei Li, Pingyu Cai, Nian Li designed the study. Jinxia Liu, Yaoxin Ruan, Jiaxiang Pan and Pingyu Cai extracted, collected and analyzed data. Meimei Li, Fengjing Cai, Chaoyang Xu, Huili Lin reviewed the results, interpreted data, and wrote the manuscript. All authors read and approved the final manuscript.

## Data Availability

The datasets generated and analyzed during the current study are available from the corresponding author on reasonable request.

## Consent for publication

Not applicable.

## Ethics statement

This study was conducted in accordance with the principles of the Helsinki Declaration. The MIMIC-IV database ensures patient privacy by de-identifying all personal information and assigning random codes for patient identification. Given the retrospective design and the use of anonymized data, the Ethics Committee of the Second Affiliated Hospital of Fujian Medical University granted a waiver for informed consent.

---

**sFigure 1.**
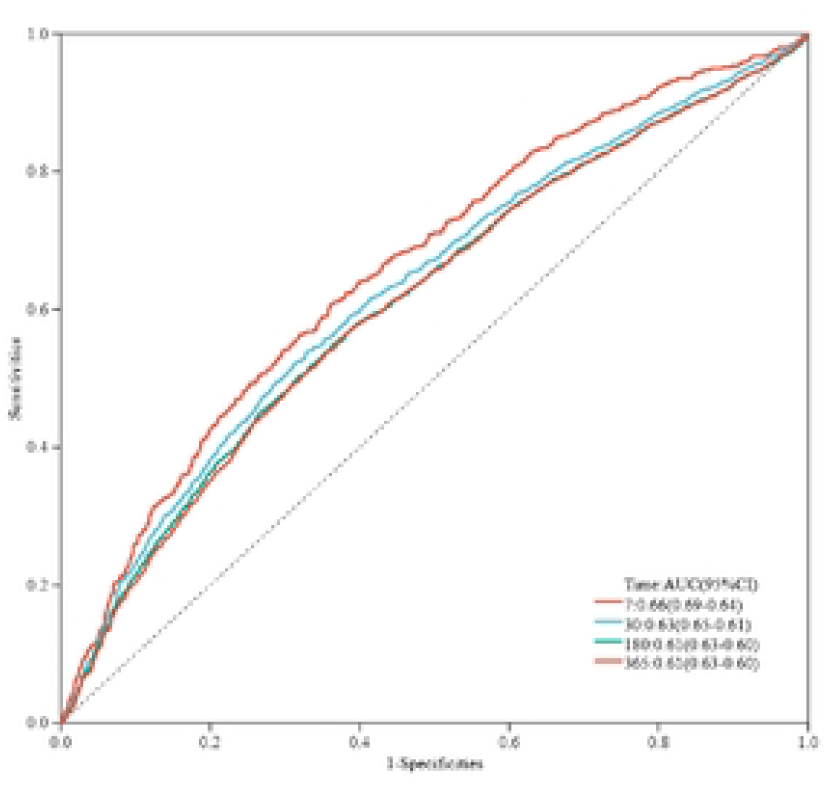
Receiver operating characteristic Curves for Predicting 7-Day, 30-Day, ISO-Day and 365-Day Mortality.

